# A Phase II Study Repurposing Atomoxetine for Neuroprotection in Mild Cognitive Impairment

**DOI:** 10.1101/2021.07.06.21260104

**Authors:** Allan I. Levey, Deqiang Qiu, Liping Zhao, William T. Hu, Duc M. Duong, Lenora Higginbotham, Eric B. Dammer, Nicholas T. Seyfried, Thomas S. Wingo, Chadwick M. Hales, Malú Gámez Tansey, David Goldstein, Anees Abrol, Vince D. Calhoun, Felicia C. Goldstein, Ihab Hajjar, Anne M. Fagan, Doug Galasko, Steven D. Edland, John Hanfelt, James J. Lah, David Weinshenker

## Abstract

The locus coeruleus (LC) is the initial site of Alzheimer’s disease neuropathology, with hyperphosphorylated Tau appearing in early adulthood followed by neurodegeneration in dementia. LC dysfunction contributes to Alzheimer’s pathobiology in experimental models, which can be rescued by increasing norepinephrine (NE) transmission. To test NE augmentation as a potential disease-modifying therapy, we performed a biomarker-driven phase II trial of atomoxetine, a clinically-approved NE transporter inhibitor, in subjects with mild cognitive impairment due to Alzheimer’s disease.

The design was a single-center, 12-month double-blind crossover trial. Thirty-nine participants with mild cognitive impairment (MCI) and biomarker evidence of Alzheimer’s disease were randomized to atomoxetine or placebo treatment. Assessments were collected at baseline, 6- (crossover) and 12-months (completer). Target engagement was assessed by CSF and plasma measures of NE and metabolites. Prespecified primary outcomes were CSF levels of IL1α and Thymus-Expressed Chemokine. Secondary/exploratory outcomes included clinical measures, CSF analyses of Aβ42, Tau, and pTau_181_, mass spectrometry proteomics, and immune-based targeted inflammation-related cytokines, as well as brain imaging with MRI and FDG-PET.

Baseline demographic and clinical measures were similar across trial arms. Dropout rates were 5.1% for atomoxetine and 2.7% for placebo, with no significant differences in adverse events. Atomoxetine robustly increased plasma and CSF NE levels. IL-1α and Thymus-Expressed Chemokine were not measurable in most samples. There were no significant treatment effects on cognition and clinical outcomes, as expected given the short trial duration. Atomoxetine was associated with a significant reduction in CSF Tau and pTau_181_ compared to placebo, but not associated with change in Aβ42. Atomoxetine treatment also significantly altered CSF abundances of protein panels linked to brain pathophysiologies, including synaptic, metabolism, and glial immunity, as well as inflammation-related CDCP1, CD244, TWEAK, and OPG proteins. Treatment was also associated with significantly increased BDNF and reduced triglycerides in plasma. Resting state fMRI showed significantly increased inter-network connectivity due to atomoxetine between the insula and the hippocampus. FDG-PET showed atomoxetine-associated increased uptake in hippocampus, parahippocampal gyrus, middle temporal pole, inferior temporal gyrus, and fusiform gyrus, with carry-over effects six months after treatment.

In summary, atomoxetine treatment was safe, well tolerated, and achieved target engagement in prodromal Alzheimer’s disease. Atomoxetine significantly reduced CSF Tau and pTau, normalized CSF protein biomarker panels linked to synaptic function, brain metabolism, and glial immunity, and increased brain activity and metabolism in key temporal lobe circuits. Further study of atomoxetine is warranted for repurposing the drug to slow Alzheimer’s disease progression.

## Introduction

Alzheimer’s disease is a devastating progressive dementia with tremendous societal burden, yet no disease-modifying treatments exist. With the projected dramatic age-related increasing prevalence in the next few decades, and growing numbers of failed clinical trials targeting amyloid, there is an urgent need to expand the scope of potential therapeutic targets. Genetic, epidemiological, and experimental studies have identified a multitude of risk factors that appear to converge on several biological pathways downstream of Aβ such as neurofibrillary tangle formation and neuroinflammation that damage neural circuits and synaptic transmission involved in memory, cognition, and behavior, and relentlessly drive progressive neurodegeneration.^1^ Given that Aβ deposition begins two or more decades prior to symptom onset, these downstream pathways provide new treatment targets for disease modification if initiated prior to significant neurodegeneration and dementia.

The locus coeruleus (LC), the major brainstem noradrenergic nucleus that innervates and supplies norepinephrine (NE) to the forebrain to regulate arousal, cognition, and behavior, has garnered interest in its potential as a disease-modifying therapeutic target for Alzheimer’s disease.^2^ While degeneration of the LC has long been known as a ubiquitous feature of Alzheimer’s disease,^3-7^ studies provide several lines of compelling evidence that impaired LC function in Alzheimer’s disease contributes to not only the clinical symptoms, but also triggers underlying pathophysiological mechanisms involved in progressive neurodegeneration.^2,8-20^ Both imaging and postmortem studies indicate that volumetric reduction, neuronal loss, and neuropathology in LC predict the rate of cognitive decline, attentional and executive function deficits, and Tau burden in humans, suggesting an important role in cognitive resilience and abnormal protein aggregation.^16,21-25^ Hyperphosphorylated Tau, a “pretangle” form of the protein prone to aggregation, appears in the LC before any other area of the brain, and is now considered the earliest detectable Alzheimer’s disease-like neuropathology, evident even in young and middle-aged adults.^14,26-32^ The connectivity of the LC provides a neuroanatomical substrate that may mediate the spread of pathological Tau seeds to the forebrain.^20,33^ The appearance of Tau pathology in the LC is also associated with depression and sleep disturbances, important risk factors for Alzheimer’s disease,^7,34^ and cognitive impairment becomes evident as LC neurons start to degenerate.^19^ Causal relationships between the LC and disease-modifying processes are implicated using genetic and neurotoxin-induced lesions of the LC, which exacerbate neuropathology and cognitive deficits in both amyloid- and Tau-based transgenic mouse models of Alzheimer’s disease, at least in part mediated by the critical role of LC in regulation of neuroinflammation.^2,7,8,35,36^ NE has potent effects on inflammation in the brain, where it suppresses the production and release of pro-inflammatory molecules in favor of anti-inflammatory cascades^2,8,10,11,18,37,38^ and stimulates microglial clearance of amyloid.^38^ Moreover, lesions of the LC in Alzheimer’s disease mouse model systems mirror several other features of the human disease, including regional hypometabolism, neurotrophin deficiency, blood-brain barrier permeability, and neurodegeneration.^11,15,39-41^ Finally, cutting-edge technologies that directly manipulate LC activity, such as DREADD (Designer Receptor Exclusively Activated by Designer Drugs) chemogenetics or more traditional pharmacological augmentation of NE neurotransmission, reverse the pro-inflammatory and other pathophysiological features of Alzheimer’s disease, increase microglial phagocytosis and amyloid clearance, and rescue cognitive and behavioral deficits.^9,13,38,42,43^ Compared to other therapeutic strategies, one advantage of targeting the LC-NE system is the abundance of available drugs that regulate various steps in NE transmission, from synthesis to release/reuptake and downstream receptor signaling, which have shown efficacy in cell culture and animal models of Alzheimer’s disease.^9,13,38,41,42,44-48^

To test proof of concept for NE augmentation as a potential disease-modifying therapy in humans, we initiated a biomarker-driven phase II trial of atomoxetine in mild cognitive impairment (MCI). We chose atomoxetine, a selective NE reuptake inhibitor for several reasons. The drug blocks the plasma membrane NE transporter (NET), but not other monoamine transporters,^49^ resulting in increased extracellular NE in the periphery and brain.^50,51^ Atomoxetine (in combination with the synthetic NE precursor L-3,4-dihydroxyphenylserine; L-DOPS) ameliorates glial activation and Aβ deposition, increases neurotrophin expression, and reverses cognitive deficits in a mouse model of Alzheimer’s disease.^42^ Atomoxetine also improves the phasic-to-tonic ratio of LC firing, which is associated with focused attention important for some aspects of learning and memory.^52^ It is possible to quantitatively demonstrate target engagement by measuring levels of NE and its primary metabolite 3,4-dihydroxyphenylglycol (DHPG) in blood and cerebrospinal fluid (CSF).^53^ In addition, as an FDA-approved drug widely used for treating attention disorders,^54-56^ atomoxetine is safe for chronic use in children and adults, including geriatric populations,^57,58^ and improves cognitive function in Parkinson’s disease patients with lower LC volume,^59^ providing an excellent opportunity to repurpose this medication for Alzheimer’s disease.

Here we report the results of a single-center, phase II randomized, double-blind, placebo-controlled, 6-month crossover trial. Thirty-nine subjects with MCI and biomarker results consistent with AD were randomized to atomoxetine or placebo treatment for 6 months, and then crossed over to receive the alternative intervention for 6 months. The primary outcomes of the study were safety and tolerability, CSF biomarkers of target engagement (NE metabolites), and neuroinflammation. IL1-⍰ and Thymus Expressed Chemokine (TECK, aka C-C Motif Chemokine Ligand 25 or CCL25) were preselected as primary CSF outcome markers of neuroinflammation because they best predicted subsequent cognitive decline in a preliminary study.^60^ Neither markers were not detectable in the majority of subjects in the current study, and to address this limitation, we assessed if there was a difference in non-detection between the treatment groups. Given the diversity of mechanisms by which NE augmentation can modify the neurobiology of disease in preclinical studies, a key goal of this study was to investigate the effects of atomoxetine treatment on a wide range of pathophysiological processes in addition to clinical outcomes. As such, our secondary and exploratory outcomes broadly explored a range of biomarkers using advanced proteomics and imaging methods to inform both disease biology and future clinical trial design. In addition to clinical findings and biomarkers of Alzheimer’s disease progression with CSF amyloid beta 42 (Aβ42), total Tau and phospho-Tau (pTau_181_), we used recently developed mass spectrometry (MS) methods to assess five panels of neuropathology-based protein biomarkers linked to synaptic dysfunction, glial immunity, metabolism, myelin injury, and vascular biology.^61,62^ We also used immunoassays to explore the effects of atomoxetine on cytokines and a panel of inflammation analytes,^63,64^ CSF brain-derived neurotrophic factor (BDNF), and brain imaging using volumetric MRI (vMRI), resting state functional MRI (rsfMRI), and fluorodeoxyglucose positron emission tomography (FDG-PET).

## Materials and methods

### Trial population

Eligible participants were aged 50-90, diagnosed with MCI due to Alzheimer’s disease at Emory University after comprehensive clinical assessments with board certified cognitive neurologists, and consented and enrolled between 2012 and 2018. MCI was defined as: (1) presence of subjective memory concerns; (2) meets ADNI criteria for diagnosis of amnestic MCI, either single or multidomain subtype^65^; (3) CSF levels of Aβ42, total Tau, and pTau_181_ consistent with underlying Alzheimer’s disease pathology according to established threshold values at Emory and the ADNI Biomarker Core. Other key inclusion criteria included Mini-Mental State Exam score between 24 and 30 (inclusive); Clinical Dementia Rating = 0.5 (Memory Box score at least 0.5); and Geriatric Depression Score ≤ 6. Cholinesterase inhibitors and memantine were allowable if stable for 12 weeks prior to screen. Key exclusion criteria included any significant neurologic disease other than MCI and suspected incipient Alzheimer’s disease, contraindication to MRI or MRI findings suggestive of memory loss due primarily to cerebrovascular or other structural condition, major depression or presence of suicide risk based on structured clinician interview, current use of antidepressant medications that act on NET (duloxetine, venlafaxine, desvenlafaxine, imipramine, or amitriptyline), and serious cardiac abnormalities.

### Trial oversight

The trial was registered with ClinicalTrial.gov (NCT01522404) after the protocol and informed consent form were reviewed and approved by the Emory Institutional Review Board (IRB) committee (IRB00054397). All study participants provided written informed consent and participation was in accordance with the principles of the Declaration of Helsinki. In those who lacked decision capacity, a study surrogate or study partner who could provide consent on the participant’s behalf was required. Atomoxetine and placebo were purchased by, compounded, and dispensed by Emory University Investigational Drug Services, using matched capsules for atomoxetine and placebo. The Alzheimer Disease Cooperative Study (ADCS) at UCSD provided data coordination and independent statistical analyses (Dr. Steven Edland). An independent study monitor performed periodic reviews to ensure data quality. From the initiation of the trial until the final complete analysis was conducted, all investigators were blinded to the group assignment. A local study biostatistician (LZ) presented the study data to an independent Data and Safety Monitoring Board (DSMB). The study was funded by grants from philanthropy and the Alzheimer Drug Discovery Foundation (ADDF). The ADDF influenced the design by recommending the cross-over trial to increase statistical power, but otherwise had no influence on conduct or data collection and analysis, nor on the preparation of the final manuscript or publication decision.

### Trial design

The trial was an investigator-initiated, single-center, double-blind, placebo-controlled, randomized cross-over study in the Atlanta metropolitan area, comparing the effects of oral atomoxetine vs. placebo treatment for six months. Participants underwent screening and baseline assessments, and then were randomly assigned to treatment with placebo or flexible doses of the NET inhibitor atomoxetine, starting with 10 mg po daily and increasing weekly by increments (18 mg week 2, 40 mg week 3, 60 mg week 4, 80 mg week 5) to a maximum of 100 mg po daily or the maximum tolerated dose. The local biostatistician provided a computer-generated random number to the ADCS to stratify and balance treatment and placebo arms based on APOE4 carrier and non-carrier status. Weekly phone visits during this dose escalation phase provided additional review of safety and tolerability. At the six-month time point (visit 14/week 29), baseline measures were reassessed (without a drug washout), and then subjects who were assigned to active treatment crossed over to placebo, and those subjects who were initially randomized to placebo received active treatment with atomoxetine.

### Trial procedures

Following informed consent, screening included Mini Mental Status Exam (MMSE), Logical Memory (LM) and Clinical Dementia Rating-Sum of Boxes (CDR-SB) assessments, Geriatric Depression Scale (GDS), and, if findings were consistent with MCI, additional medical history, physical and neurological exam, vital signs, electrocardiogram, Modified Rosen Hachinski, suicidality assessment, urinalysis, and blood work that included comprehensive blood count and metabolic panel, thyroid stimulating hormone, vitamin B12, coagulation panel, and CYP2D6. Baseline, 6-month and 12-month evaluations included safety measures (see below), neuropsychological testing, venous blood draws, lumbar puncture for CSF Aβ_42_, Tau, and pTau_181_, brain MRI, and FDG-PET. All participants received neuropsychological tests assessing premorbid verbal intellectual functioning (American National Adult Reading Test)^66^ and overall cognitive status (Montreal Cognitive Assessment^67^; Alzheimer’s Disease Assessment Scale-Cognitive Subscale (ADAS-Cog 13 with delayed recall and Number Cancellation)^68^). In addition, in depth measures of semantic memory (Animal Fluency)^69^, episodic memory (Auditory Verbal Learning Test) <u>70</u>, executive functioning (Trails A & B,^71^ Clock Drawing^72^), and language (30 item Boston Naming Test)^73^ were administered. Neuropsychiatric symptoms were assessed (Neuropsychiatric Inventory)^74^, and functional status and levels of independence were measured by the Activities of Daily Living scale (FAQ).^75^ Plasma atomoxetine drug levels were assessed by MS. Plasma and CSF catecholamine and NE metabolite levels were assayed by high performance liquid chromatography (Dr. David Goldstein, NINDS). CSF Aβ_42_, Tau, and pTau_181_ were assayed by on the Fujirebio Lumipulse platform (Akesogen and Dr. Anne Fagan, Washington University). MS with tandem mass tagging (TMT) was used to quantify five panels of proteins in the CSF that are strongly associated with brain neuropathology, using methods previously described.^62^ Other exploratory measures included antibody-based proteomics assays of a 92-protein inflammatory biomarker panel (Olink)^63^, as well as various cytokines, inflammation and lipoprotein associated oxidative stress biomarkers, and BDNF (labs of Dr. William Hu, Malu Tansey, and Ahn Le).

MRI scanning was performed on a 3T MRI scanner (Magnetom Prisma, Siemens, Erlangen, Germany) using a 20-channel head coil at baseline and 12 months. The MRI sequences collected include a 3D T1-weighted (T1w) magnetization-prepared rapid acquistion with gradient echo (MPRAGE) (FOV = 256mm, TR/TE/TI =2300/900/2.96ms, 1x1x1mm^3^ resolution), a 2D T2-weighted Fluid Attenuated Inversion Recovery (FLAIR), and rsfMRI (FOV=220mm, matrix size = 64, 35 slices with 4mm slice thickness and no gap, TR/TE= 2100/30ms, 200 volumes for a duration of 7 mins). The volumetric measures of the hippocampus and other cortical/subcortical regions were extracted using FreeSurfer software (version 5.3, Massachusetts General Hospital, MA, USA, https://surfer.nmr.mgh.harvard.edu) from T1w MPRAGE structural image. The rsfMRI data were preprocessed using a standard pipeline based on the statistical parametric mapping software (SPM, version 12, University College London, London, UK). Rigid body motion correction was performed using INRIalign toolbox in SPM to correct subject head motion, followed by the slice-timing correction to account for the slice acquisition timing differences. The realigned rsfMRI data were subsequently normalized using SPM’s TPM template, resampled to 3 × 3 × 3 mm^3^ isotropic voxels and further smoothed using a Gaussian kernel with a full width at half maximum (FWHM) = 6 mm. Constrained group independent component analysis^76^ as performed on the preprocessed rsfMRI data to identify 53 functional connectivity networks^77^, and the inter-network connectivity was then calculated using Pearson’s correlation coefficient between each pair of the identified networks.

FDG-PET imaging procedures followed the ADNI standard protocol.^78^ 5mCi of tracer was administered and each participant was asked to remain still and keep awake in an uptake waiting room where the ambient light was controlled to a level of twilight. After 20-min of tracer incorporation period, the participant was instructed to use the restroom and empty the bladder. Six repetitions of 5-min frames were acquired. Transmission scan was acquired following the emission scan for attenuation correction. Image reconstruction was performed using an ordinary Poisson-ordered subset expectation maximization algorithm using 6 iterations and 16 subsets. The attenuation corrected images were motion-corrected and averaged, scaled with the whole-brain value and normalized to the Montreal Neurological Institute (MNI) space. The automated anatomical parcellation (AAL) template^79^ was then used to calculate the standard uptake value ratio (SUVR) for each brain region defined in the template.

### Safety

Safety assessments included a physical and neurologic examination prior to randomization, periodic assessment of medical history data and clinical measures, suicidality assessment, blood pressure, heart rate, routine laboratory measurements, electrocardiogram, and surveillance of adverse events. Significant events identified during the study period through self-report by the participant or next of kin or clinically significant abnormal laboratory results were recorded as adverse events. Participants were screened for a new diagnosis of dementia at each visit.

### Statistical analysis

Data were analyzed by the intention-to-treat principle. Participants’ demographic and clinical characteristics at baseline were compared between the two treatment sequences using two-sample t-tests or chi-square tests. Baseline cognitive scores were compared with the nonparametric Wilcoxon-Mann-Whitney test between the two sequences because the scores were not normally distributed. Adverse events and dropout rates were compared between patients receiving atomoxetine and placebo via Fisher’s exact test.

To examine the effect of atomoxetine on CSF and plasma biomarker level, we log-transformed the data to adjust for skewness and conducted repeated-measures linear regression analyses under a compound symmetry covariance model. We exponentiated the fitted regression coefficients so that the results were interpretable as the percent change in biomarker abundance at crossover or final visit compared to baseline. In a sensitivity analysis, we also re-fitted the repeated measures model under an unstructured covariance structure and the results were similar (data not shown). All regression models of the treatment effect were adjusted for crossover design by including sequence, period, and baseline value effects.

Because CSF inflammation biomarker CD244 had several values below the limit of detection, for this particular biomarker we adjusted for possible bias attributable to left censoring by using a nonlinear repeated-measures model that explicitly incorporated the left censored observations in the maximum likelihood estimation. Two other biomarkers, namely the neuroinflammation markers IL1-⍰ and CCL25, were detected in only 15.1% and 49.1% of CSF samples, respectively. To address this limitation, we dichotomized these data by their detectability and used repeated-measures logistic regression analyses, adjusted for the crossover design, to assess if there was a difference in the odds of non-detection between atomoxetine and placebo.

Cognitive variables were analyzed in a manner similar to the other biomarkers, but we did not first log-transform the cognitive data before conducting repeated-measures linear regression analysis.

The dose-response effect of atomoxetine concentration on percent change in the biomarkers and cognition was evaluated using Pearson correlation during the active treatment period.

The validity of the above statistical approaches for cross-over designs was sensitive to the assumption that there was no sustained effect of atomoxetine six months after treatment was stopped. To test this assumption, we investigated whether there were carry-over effects using Grizzle’s method for crossover trials,^80^ as well as a refinement of Grizzle’s method that adjusted for baseline value using linear regression. In particular, we found evidence of a carry-over effect in the FDG-PET biomarkers; as these particular biomarkers were normally distributed, the effect of atomoxetine on FDG-PET biomarkers was determined by an unadjusted two-sample t-test based on the first treatment period only.

All tests performed were two-tailed. In this exploratory study, unless indicated otherwise we did not adjust for multiple comparisons: p-values less than 0.05 were regarded as significant. As we indicate, an exception is that for cognitive outcomes we declared results significant if the false discovery rate was less than 5%.^81^ All analyses were carried out in SAS 9.4, and Matlab with SPM software.

CSF protein panels were assembled from selected proteins corresponding to panels defined in Higginbotham et al.^61^ as protein isoforms with less than 50 percent missing quantified normalized abundance by Proteome Discoverer software (Thermo Scientific). Normalized protein abundances across 14 TMT batches were batch-corrected using a robust median polish^62^ of abundance, followed by calculation of individual protein log_2_ abundance ratios for post-6 month ATX/placebo or baseline (N=36; or placebo/baseline, N=19) for paired longitudinal CSF samples. Then, a Z-score transformation was applied to the log_2_ ratios of proteins that corresponded to the five panels, and the average Z score representing panel abundance within each patient was calculated. Finally, a T test was used to check for significance of change between the placebo/baseline and ATX/(placebo or baseline) Z scores. Panels were comprised of measurements for the metabolic protein panel (KRT2, PKM, ALDOA, ENO2, GOT1, PGK1, PTI1, GPI, GOT2, MDH1, PGAM1, YWHAG, and CALM2); the synaptic protein panel (BASP1, GDA, GAP43, YWHAZ, LDHA, HK1, AP2B1, YWHAB, HPRT1, and DTD1); the glial protein panel (ALDOC, ENO1, SPON1, MARCKS, PARK7, SMOC1, GLOD4, GMFB, and GLO1); the myelin protein panel (SPP1, 4 isoforms, PTPRZ1, SOD1, GDI1, PEBP1, DDAH1, PPIA, GSS, and GSTO1); and the vascular protein panel (CP, 2 isoforms, F2, KNG1, 2 isoforms, COL6A1, C9, NID2, AMBP, AHSG, VTN, OGN, LAMA5, LUM, PON1, AEBP1, MFGE8, COL14A1, OLFML3, DCN, and NUCB2).

## Data Availability

The data that support the findings of this study are available from the corresponding author, upon reasonable request.

## Results

After screening 56 individuals, 39 participants were enrolled in the trial, and 36 completed the study as shown in the flow chart (Figure 1). Two of the participants withdrew during the atomoxetine treatment period, and one withdrew while on placebo. One participant withdrew after crossover following diagnosis with cancer and was not able to finish testing.

**Figure 1.**
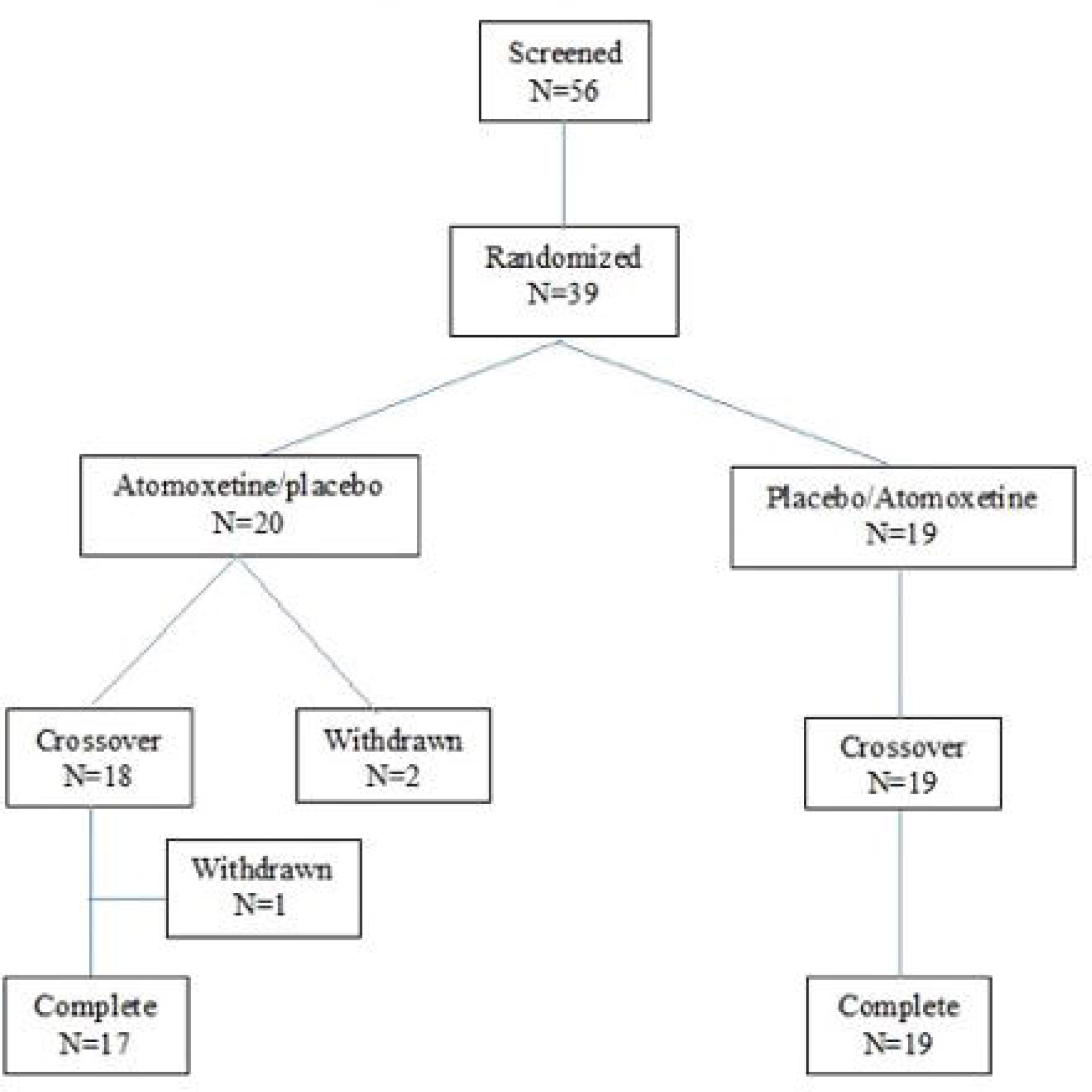
Atomoxetine study participant flow chart.

### Baseline Characteristics

Participant demographic, clinical characteristics and cognitive scores were similar at baseline between the two randomization arms (Table 1).

### Primary Outcomes

#### Safety and Tolerability

The primary outcome of safety and tolerability was predesignated as a dropout rate < 15%. The dropout rate was 5.1% (2 of 39) in the subjects treated with atomoxetine and 2.7% (1 of 37) in those treated with placebo (p=0.99). All but two subjects who completed the trial were successfully titrated to the maximum dose of 100 mg. There were no significant differences in serious adverse events between treatments, with 3 (7.7%) occurring with atomoxetine (two of which may have been related to study drug; dizziness and dysautonomia) and 3 (8.1%) with placebo (p=0.99).

The total number of adverse events was higher when treated with atomoxetine compared to placebo (Table 2). In relation to study drug, there were 145 adverse events reported with atomoxetine treatment and 88 with placebo, including some considered definitely related (5 atomoxetine; 0 placebo), and possibly related (47 atomoxetine; 17 placebo). The most common adverse events associated with atomoxetine treatment (Supplementary Table 1) were gastrointestinal symptoms (12 atomoxetine; 4 placebo), dry mouth (10 atomoxetine; 2 placebo), and dizziness (10 atomoxetine; 8 placebo).

Heart rate increased ∼8-9 beats per minute (bpm) on atomoxetine (unadjusted analysis, Supplementary Table 2). Repeated measures analysis with adjustment of baseline measures for sequence, period, and treatment also showed that atomoxetine treatment increased heart rate (5.4 +/-2.4 bpm, p=0.029, and 9.2% +/- 3.5 from baseline). Blood pressure did not change significantly while subjects received atomoxetine compared to placebo, although there was a trend for elevated diastolic blood pressure. Body weight decreased about 4 lb on atomoxetine (unadjusted analysis, Supplementary Table 2), which after adjustment for baseline corresponds to 2.4% loss of weight (CI: -3.5, -1.3; p=0.0001).

### Compliance and Target Engagement

Compliance was excellent as verified by assessments of plasma levels of atomoxetine throughout the trial. All participants had detectable atomoxetine during the active treatment period at each visit, with the exception that drug levels were undetectable only in one visit each for 3 of 37 participants (1 of whom withdrew), and these were all at the penultimate visit before crossover or completion. Once subjects had achieved highest dose after titration, the median atomoxetine plasma concentrations were 313.8 ng/ml (110.6 – 701, interquartile range) for the active/placebo arm, and 224.4 ng/ml (56.9 - 536.4) for the placebo/active arm. The major active metabolite, 4OH-atomoxetine, was also similar in all subjects during the active treatment period (8.5 ng/ml [3.7 – 14.2], active/placebo arm; 7.4 ng/ml [3.8 – 14.7] placebo/active arm, respectively).

Since the primary mechanism of action of atomoxetine is inhibition of the NET, we first measured target engagement by measurements of CSF NE and DA, both of which are substrates for the NET. As shown in Figure 2, atomoxetine significantly increased NE and DA (p<0.0001), and reduced the catecholamine metabolites DHPG (p<0.0001), DOPAC (p<0.05), and cysteinyl DA (p<0.05). There was a trend for a reduction of EPI, but it did not reach significance (p=0.05), and there was no effect on the catecholamine precursors DOPA and cysteinyl DOPA (p>0.7). Similarly, atomoxetine significantly increased plasma NE and DA levels, and reduced DHPG, DOPA, DOPAC and cysteinyl DOPA (data not shown). Hence, atomoxetine successfully achieved robust target engagement with inhibition of NET in both brain and the periphery.

**Figure 2.**
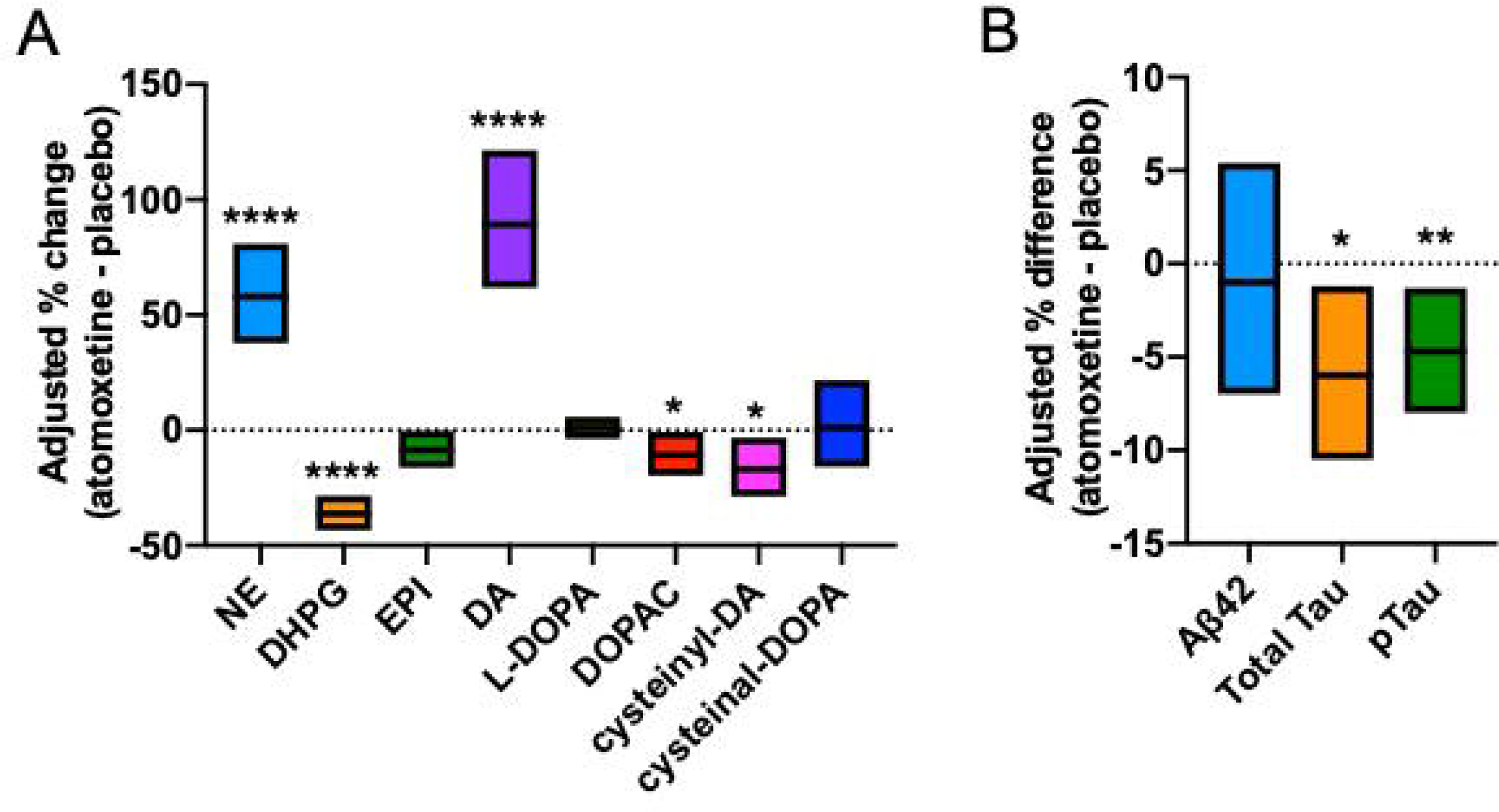
Adjusted Effect of Atomoxetine vs Placebo on CSF Biomarkers of Target Engagement and AD. (**A**) Catecholamine biomarkers show target engagement with norepinephrine (NE) reuptake inhibition, increasing levels of NE and dopamine (DA), both substrates for the NE transporter. Values are estimated differences and corresponding 95% confidence (n=36). (**B**) Alzheimer’s disease biomarkers Aβ42, Tau, pTau_181_ assays for were performed using the using Automated Lumipulse G System (Fujirebio) and analyzed by logistic regression of change in Z-score after adjustment for baseline values. Atomoxetine treatment compared to placebo significantly reduced CSF levels of total tau (n=33) and pTau_181_ (n=36) and had no effect on CSF Aβ42 levels (n=34). * p<0.05; **p<0.01: ****p<0.0001

### CSF Biomarkers of Neuroinflammation

IL1-⍰ and TECK were preselected as primary CSF outcome markers of neuroinflammation because they best predicted subsequent cognitive decline in a preliminary study in MCI^60^. However, only 15.1 % of the CSF samples were above the limit of detection for IL1-⍰, and 49% of the samples for TECK, precluding the planned analysis of comparing mean levels of these analytes across treatment groups. To address this limitation, we compared their detectability in a dichotomous manner. There was a trend for atomoxetine treatment to suppress CSF IL-1α levels below the limits of detection. After adjustments for possible baseline, period, and sequence effects, the odds ratio of having non-detectable CSF IL-1α for atomoxetine treatment versus placebo was 2.7 (CI: 0.8 - 8.9, p value=0.10). There was no difference between the treatments in the odds of having undetected TECK (OR 1.31, CI: 0.6-3.1, p=0.54).

## Secondary and Exploratory Outcomes

Other potential measures of disease modification were explored with the goal of informing a subsequent larger and longer trial focused on the potential disease-modifying properties of NE signaling. For this purpose, a broad array of measures of disease progression and neurodegeneration (CSF Alzheimer’s disease biomarkers Aβ42, Tau and pTau_181_, vMRI, rsfMRI, and FDG-PET) were determined, as well as other biofluid markers of inflammation, neurotrophin signaling, and oxidative stress that have also been linked to NE neurotransmission in preclinical models.

### Clinical outcomes

There was no significant difference (p = 0.31) in the proportion of participants who converted from MCI to dementia between the periods when subjects were treated with atomoxetine (3/39) versus placebo (6/37), as expected for only 6 months of treatment. There was also no treatment effect on general measures of cognition with MMSE (-0.6%; CI: -4.6%, 3.3%) and MoCA (- 2.5%; CI: -6.9%, 1.8%). Treatment of atomoxetine did not significantly alter performance on other neuropsychological measures with the exception of slight worsening on the ADAS-13 and Trails B; with the differences non-significant after false discovery rate (FDR) adjustment (Supplementary Figure 1).

### CSF Alzheimer Disease Biomarkers

Aβ42, Tau, and pTau_181_ assays were performed using the Automated Lumipulse G System and analyzed by repeated measure analysis after adjustment for baseline values, sequence and period. The biomarkers were log-transformed in the model. Atomoxetine treatment significantly reduced CSF levels of total Tau by 6% (CI: 1.4%, 10.3%, p = 0.017) and pTau_181_ by 4.7% (CI: 1.5%, 7.9%, p=0.008) compared to placebo, and had no effect on CSF Aβ42 levels (-1.0%, CI: -6.7%, 5.1; p=0.75) (Figure 2B). Because of the cross-over trial design, we also examined the possibility of carry-over effects where active drug administration in the first 6 months might have longer term effects on the outcomes after placebo treatment at 12 months. We found no evidence for carry-over effects on CSF Aβ42, Tau, and pTau_181_ using Grizzle’s statistical method, and a refinement of Grizzle’s method that uses linear regression to adjust for baseline values.

### Mass Spectrometry-Based CSF Biomarker Panels Linked to AD Brain Pathophysiologies

We recently developed a novel MS approach to quantify five panels of proteins in the CSF that are strongly associated with brain neuropathology.^61,62^ These panels reflect clusters of 60 proteins that are co-expressed and significantly altered in both Alzheimer’s disease brain and CSF, and importantly, they reflect distinct physiological and pathophysiological processes including synaptic function, glial immunity, metabolism, myelination and vascular biology.^61^ Using our targeted MS approach, we investigated the effect of atomoxetine treatment on the abundance of 60 proteins that comprise the respective biomarker panels. Using a TMT-MS approach, we measured the CSF abundance levels of these five panels (Z-score) in each of the 36 subjects at baseline, 6 months, and 12 months. The ratio of abundance levels before and after treatment with either placebo or atomoxetine therapy were calculated for each arm of the trial. These ratios were then used to investigate the effect of atomoxetine treatment on each of these biomarker panels compared to placebo. Measurements of placebo effect were derived only from the placebo/active arm due to concern that placebo responses in the active/placebo arm may be confounded by carryover effects from earlier atomoxetine treatment. The results demonstrated significant decreases in the synaptic and metabolic panels following atomoxetine treatment compared to placebo (Figure 3A). These neuron-associated panels are increased in Alzheimer’s disease CSF compared to controls,^61^ indicating that atomoxetine exerted a normalizing effect on these biomarkers. The glial immunity panel, which is similarly elevated in Alzheimer’s disease compared to controls,^61^ also demonstrated notably decreased levels following atomoxetine treatment that trended toward significance. An analysis of alterations in individual panel proteins revealed primarily synaptic (e.g., LDHA, YWHAB) and metabolic (e.g., PGAM1) among those most significantly decreased following atomoxetine therapy (Figure 3B). The inflammation-associated protein ENO1, as well as DDAH1 of the myelin panel, were also highly decreased after atomoxetine. The vascular panel, which demonstrates decreased levels in Alzheimer’s disease compared to controls,^61^ yielded two markers (COL14A1, NID2) with significantly increased levels following atomoxetine therapy. Overall, these findings suggest atomoxetine has a normalizing effect on various Alzheimer’s disease CSF biomarkers across a wide range of pathophysiologies, though most notably those associated with synaptic, metabolic, and inflammatory pathways.

**Figure 3.**
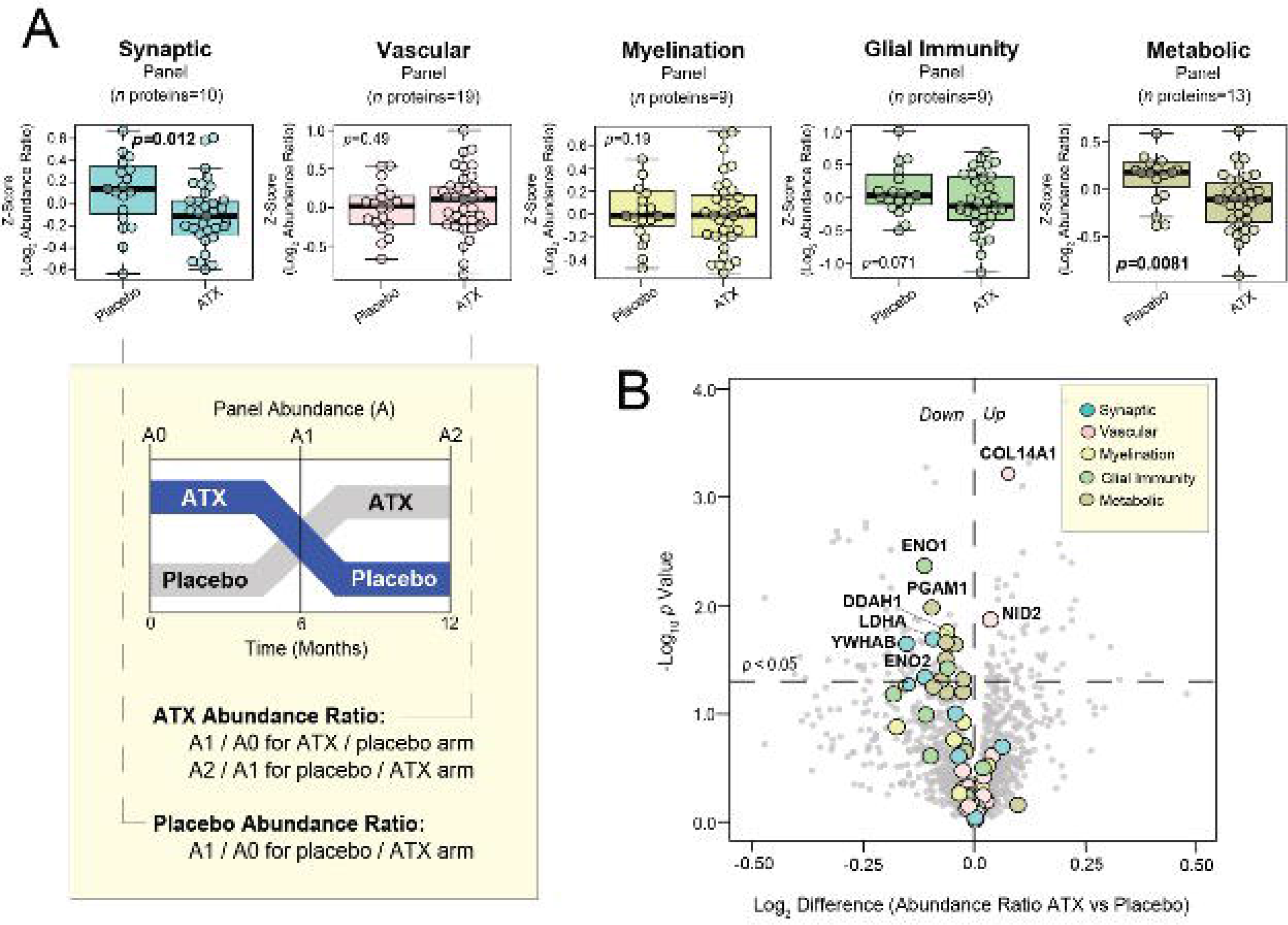
Atomoxetine therapy exerts normalizing effect on CSF AD biomarkers associated with synaptic, metabolic, and inflammatory pathophysiology. Longitudinal CSF samples collected from each of the 36 subjects at baseline, 6 months, and 12 months were analyzed by TMT-MS and Alzheimer’s disease biomarker pathway panels were quantified. For each subject, abundance ratios before and after treatment with either placebo or atomoxetine therapy were calculated for each arm of the trial. The placebo effect measurements were derived from only the placebo / active arm of the trial due to concern that post-placebo responses in the active / placebo arm may be confounded by carryover effects from earlier ATX therapy. (**A**) Box plots demonstrating the log_2_-transformed ratio of pre- and post-treatment panel abundance levels (Z-score) following 6 months of either placebo or atomoxetine (ATX) therapy. T-test analysis was used to identify panels with significantly different (*p*<0.05) post-treatment responses to placebo and ATX. (**B**) Volcano plot displaying the log_2_-transformed difference in abundance ratio (x-axis) against the -log_10_ statistical *p* value (y-axis) for all proteins demonstrating differential responses following 6 months of ATX therapy compared to placebo. Proteins belonging to biomarker pathway panels are represented by colored data points.

### Biofluid Biomarkers of Inflammation, Neurotrophin Signaling, and Oxidative Stress

In CSF, there were no significant differences between atomoxetine treatment and placebo for levels of C3, IL-6, IL-7, IL-9, IL-10, IP-10, NfL, TNF or VEGF using Luminex and Mesoscale platforms (Supplementary Figure 2A). Measures of IL-1β IL-4, IL-17A, and BDNF were under the limit of detection in all samples. Because of the detectability challenges, we also performed exploratory analyses of a panel of 92 inflammation analytes in CSF using a more sensitive, specific, and precise multiplexed proximity extension assay (Olink). On this platform, CDCP1, CD244 and TWEAK were significantly reduced (p<0.05 unadjusted), and OPG significantly increased (p<0.006) by atomoxetine treatment (Supplementary Figure 2B). We observed no carry over effects of atomoxetine on any of these analytes.

Plasma BDNF levels were significantly increased by 24.0% (CI: 1.8%, 51.0%; p=0.04, unadjusted), and triglycerides significantly reduced by -13.4% (CI: -24.5%, -0.7%; p=0.048, unadjusted) by atomoxetine. IL-1β, IL-4, C3, IL-6, IL-11, IL-17A, IP-10, and TNF showed no significant differences. Other plasma biomarkers of lipoprotein-associated oxidative stress and inflammation showed no differences with atomoxetine versus placebo treatment for cholesterol, HDLc, LDLc, oxLDL, apoAI, apoB, apoE, hsCRP, nitrotyrosine, and glutamine.

### MRI and PET Imaging Biomarkers

#### MRI

There were 101 MRI scans for 36 patients (17 in the “ATX/Placebo” and 19 in the “Placebo/ATX” arms). Unadjusted comparison of percent of change in the brain region volume at crossover and completion from baseline were compared by two-sample t-test and there was no significant change (p>0.1). Linear mixed effects models that adjusted for the crossover design were fitted. Significant difference in the percent of change between the two treatments was seen in ventral diencephalon only (p=0.016).

#### Resting state-fMRI

Static functional connectivity between the 53 Neuromark^77^ template regions was estimated to investigate treatment effects from fMRI data. A linear mixed method adjusted for baseline static functional connectivity values, age, gender and BMI was run to investigate significant treatment by time interactions, and p-values were false discovery rate (FDR) corrected for multiple comparisons at a significance level of 5%. Between these 53 identified functional connectivity networks, significantly increased inter-network connectivity due to atomoxetine treatment was found between the insula and the hippocampus, while decreased inter-network connectivity was found between the inferior frontal gyrus (IFG) and the caudate networks.

### FDG-PET

There were 106 PET scans for 36 patients (17 in the “ATX/Placebo” and 19 in the “Placebo/ATX” arms). We evaluated the effects of atomoxetine on brain metabolism on 45 regions defined on the AAL template using FDG-PET. We found interesting patterns of brain regions showing potential treatment-induced effects (Figure 4). Significantly increased glucose uptake due to atomoxetine was found in the hippocampus (p = 0.036), the parahippocampal gyrus (p = 0.023), the middle temporal pole (p = 0.021), the inferior temporal gyrus (p = 0.022) and the fusiform gyrus (0.027), whereas significantly decreased SUVR due to ATX was found in the inferior frontal orbital gyrus (p = 0.046), and the calcarine (p = 0.023). Carryover effects of atomoxetine were also significant in each of these regions using three different statistical methods, with the exception of inferior frontal orbital gyrus.

**Figure 4.**
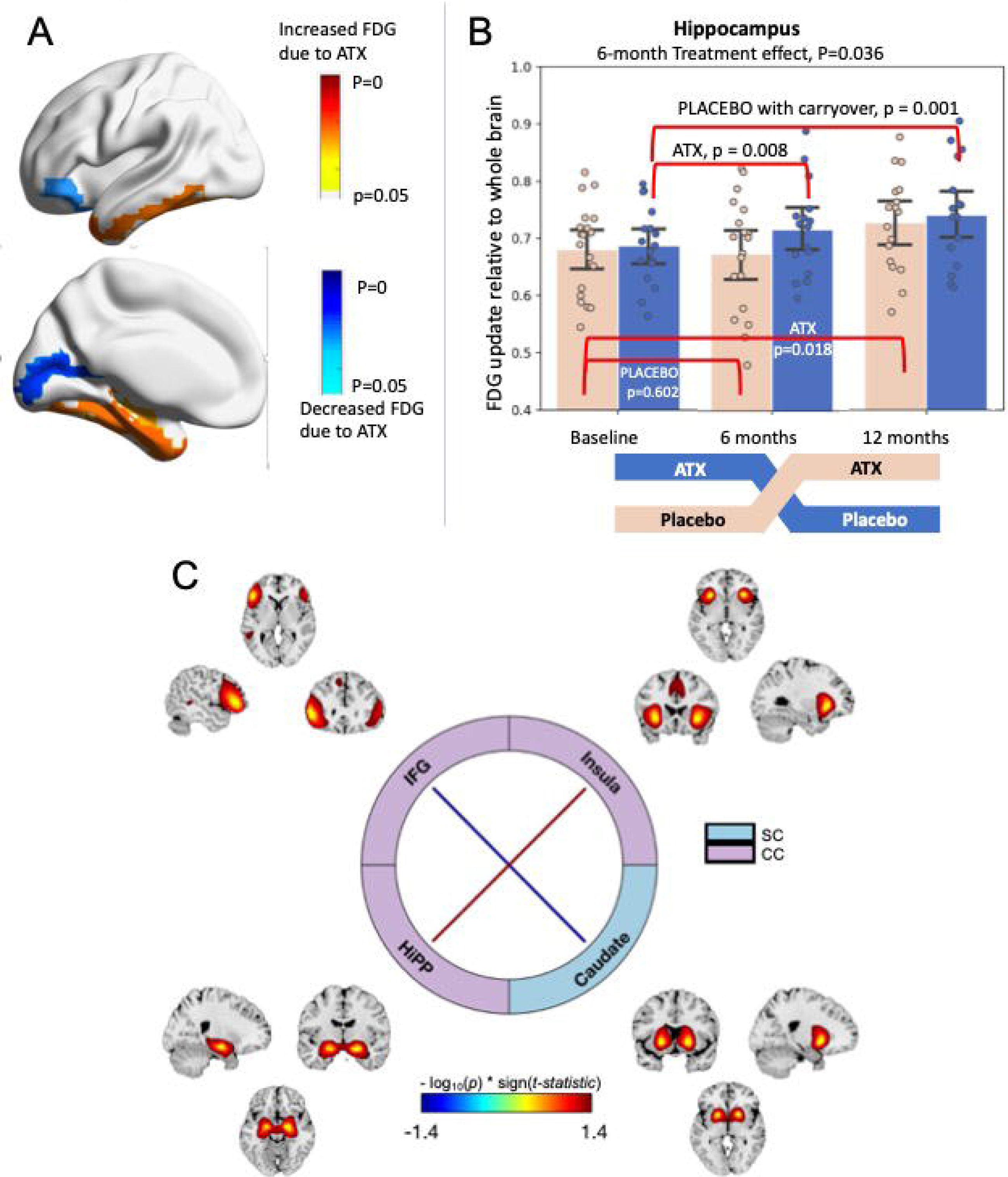
Effect of Atomoxetine vs Placebo on PET and MRI Imaging Biomarkers. (**A**) 3D rendering of regions showing significantly increased (warm colors) and decreased (cold colors) FDG uptake. (**B**) Quantitative values of SUVR in the hippocampus. (**C**) Resting state fMRI shows brain networks of inferior frontal gyrus (IFG), insula, hippocampus (Hipp), and caudate. The inter-network connectivity increased significantly between insula and Hipp due to atomoxetine treatment, and decreased significantly between IFG and caudate networks.

## Discussion

The LC plays an important role in cognition and behavior and, most recently, it has been recognized as the initial site of neuropathology in human brain and a driver of both amyloid plaque and neurofibrillary tangle progression in preclinical studies. In this phase II study of atomoxetine in subjects with MCI due to underlying Alzheimer’s disease pathology, we found the drug was safe, well tolerated, achieved target engagement and provided proof of concept for several potential disease-modifying properties. All participants showed excellent target engagement as determined by CSF catecholamine analysis. Over 6 months of treatment, there was also a small but significant 5-6% reduction in CSF levels of both total Tau and pTau_181_, providing biomarker evidence for potential slowing of neurodegeneration. Because of the pleiotropic effects of NE on disease biology, including microglial function, Aβ42 phagocytosis and clearance, Tau progression, blood brain barrier permeability, and neurotrophin signaling in preclinical models of Alzheimer’s disease, we also explored a wide range of biomarkers in the study to inform subsequent clinical trials. CSF levels of brain-based panels of synaptic and glial immunity biomarkers, as well as several inflammatory analytes in an Olink panel, showed treatment effects. Strong atomoxetine treatment effects were also seen on brain metabolism, evidenced by increases in FDG-PET in key medial temporal lobe circuits, as well as on a panel of brain-derived CSF proteins linked to metabolism.

Drug repurposing with atomoxetine offers several advantages given a wealth of clinical experience to reduce safety concerns and leverage the well-studied pharmacology to confirm target engagement. The study demonstrated that all but two subjects tolerated the maximal dose titration to 100 mg daily, with mild and tolerable side effects. These side effects were expected given the clinical experience with this FDA approved drug for attention deficit symptoms in children and adults, and included, most commonly, gastrointestinal symptoms and dry mouth.^54-56,82^ A few previous studies in elderly patients with depression or Alzheimer’s disease have similarly found drug treatment generally well tolerated.^57,83,84^ We confirmed levels of plasma atomoxetine by mass spectrometry in concentrations within the therapeutic ranges,^55,82,85^ with median atomoxetine plasma concentrations of 224.4 and 313.8 ng/ml during active treatment phase of each arm of the study. These levels are comparable to those observed in extensive metabolizers, as we expected, since we prescreened to exclude potential carriers of CYP2D6 genetic variants that are seen in about 10% of the general population who slowly metabolize atomoxetine and accumulate plasma concentrations over 700 ng/ml.^82,85^ The major active metabolite, 4OH-atomoxetine, was also similar in all subjects during the active treatment period (8.5 ng/ml [3.7 – 14.2], active/placebo arm; 7.4 ng/ml [3.8 – 14.7] placebo/active arm. Given atomoxetine’s known primary mechanism of action as a selective NET inhibitor, treatment resulted in the expected marked increases in CSF levels of NE and DA, the primary substrates for the NET. These results provided conclusive evidence for CNS target engagement in a manner that is often not possible with new therapeutic targets.

One previous randomized clinical trial tested the symptomatic effects of six months of atomoxetine treatment (25-80 mg daily) in 92 patients with mild to moderate AD,^57^ and found no clinically significant changes in the primary outcome cognitive function measured with the ADAS-Cog. We also found no symptomatic response to atomoxetine, although our study was not powered for cognitive outcomes given the slow rate of progression over 6 months in MCI. Several other small clinical studies in patients with Parkinson’s disease have reported atomoxetine improved executive function,^83,84,86,87^ whereas our study was associated with slightly worse performance on executive function. It was recently reported that atomoxetine preferentially improves response inhibition in Parkinson’s disease patients with low LC integrity (as measured by neuromelanin-sensitive MRI) compared to those with relatively preserved LCs, suggesting that atomoxetine is most beneficial for patients with impaired NE transmission.^59^ Longer term study of neuropsychological performance with atomoxetine combined with MRI measures of LC integrity will be important in future trials for MCI, as a disease-modifying effect with chronic treatment would be expected to slow the rate of cognitive decline, particularly in those individuals with low baseline noradrenergic capacity.

As a starting point for investigating the potential disease-modifying effects of atomoxetine, we used the “AT(N)” research framework to classify the hallmark Alzheimer’s disease pathologies, amyloid plaques (“A”) and neurofibrillary tangles (“T”); “(N)” denotes neurodegeneration or neural injury, and is bracketed in parentheses to denote that this subclassification is not disease specific.^88^ This schema considers CSF Aß42 or amyloid PET as measures of amyloid (A), CSF pTau_181_ or Tau PET as measures of neurofibrillary tangles (T), and either CSF total Tau, volumetric MRI, or FDG-PET as non-specific markers of neurodegeneration (N). In our study, “A” was assessed by CSF Aß42 measures which showed no change with atomoxetine treatment. By contrast, several “T” and “N” markers of tangles and neurodegeneration in CSF and imaging exhibited treatment-related effects. CSF levels of both pTau_181_ and total Tau both showed small but significant 5-6% reductions, which may be related to the ability of NE to disrupt Tau protofilaments and facilitate Tau degradation.^89,90^ With the exception of Tau immunotherapies,^91^ we are unaware of any other treatments thus far shown to alter CSF Tau or pTau_181_ levels. While the clinical significance of this relatively small degree of change after 6 months of treatment is unknown, a 10-12% annual reduction could potentially have a meaningful impact over years. Perhaps the most important biomarkers in clinical trials will be those that directly reflect neurodegeneration, such as rates of brain atrophy (e.g., cortical thickness) as measured by volumetric MRI. As expected given the slow rates of cortical atrophy,^92^ 6 months of treatment showed no changes, and the effect of atomoxetine on reduced atrophy of the ventral diencephalon is of unclear significance. FDG-PET is also considered a marker of “N”, and in unadjusted analyses atomoxetine treatment resulted in significantly increased SUVR in the hippocampus, parahippocampal gyrus, fusiform gyrus, inferior temporal gyrus and the middle temporal pole. These should be considered exploratory findings, especially since there was no cognitive improvement in our study. Nonetheless, they are quite striking for the regional selectivity of these five regions associated with the most vulnerable neural circuits in Alzheimer’s disease and linked to early neurofibrillary tangle deposition. Moreover, after adjusting for treatment sequence, we found a significant carry-over effect for those individuals receiving atomoxetine during the first 6 months of the trial, with increased FDG uptake persisting at 12 months compared to baseline (Figure 4B). Collectively, the CSF and imaging ATN biomarkers show promise for the disease-modifying potential of atomoxetine in MCI.

An important goal of the study was to investigate atomoxetine’s potential effects on a wide range of other biomarkers reflective of various pathophysiological processes implicated in Alzheimer’s disease and also linked to LC and NE functions in preclinical studies. The pleiotropic effects of LC and NE neuromodulation include roles in synaptic plasticity, inflammation, metabolism, and other brain-based physiological and pathophysiological processes that have been difficult to assess in vivo in humans. To address our goal to identify promising mechanism-based markers of disease-modification for future larger clinical trials, we used our recently developed integrative mass spectrometry-based strategy to identify novel CSF protein biomarker panels that correlate strongly with the same proteins changed in Alzheimer’s disease brain. We previously demonstrated that these biomarker panels are robust, reproducible across multiple independent cohorts consisting of hundreds of cases, and that they reflect Alzheimer’s disease-associated alterations in synaptic function, metabolism, glial immunity, vascular, and myelin biology.^61^

Here we tested for the first time the utility of these protein panels to inform treatment responses in a clinical trial. Interestingly, we observed significant effects of atomoxetine on the synaptic and metabolic panels, with a trend in the glial immunity panel. In each panel, atomoxetine effects tended to normalize protein expression in CSF, suggesting reversal of brain pathophysiologies. For example, in MCI brain there is reduced expression of proteins involved in synaptic function and metabolism,^62^ strongly anti-correlated to increased levels of the same proteins in CSF. Atomoxetine reduced CSF levels of both of these protein panels, suggesting improved synaptic function and brain metabolism as has been observed in preclinical studies with manipulations that increase NE signaling.^11,15,39-41^ In contrast, astrocyte- and microglia-linked neuroinflammatory processes (and protein panels) increase in MCI and Alzheimer’s disease brain and directly correlate with increases in the same proteins in CSF. Because atomoxetine decreased this protein panel in MCI, we infer that treatment reduced brain inflammation, consistent with its established mechanism of anti-inflammatory effects in Alzheimer’s disease animal models. ^2,8,10,11,18,37,38^ Atomoxetine increased FDG-PET uptake in temporal lobe regions, providing additional evidence that the hypometabolism typically seen in these regions in Alzheimer’s disease was improved. The effects of atomoxetine on the synaptic, metabolism, and glial immunity protein panels were specific, as there were no significant effects on the vascular or myelination protein panels. We also assessed CSF inflammatory protein changes using an independent sensitive and specific PEA assay,^63^ and found that atomoxetine significantly altered CSF levels of several other immunomodulatory proteins including CDCP1,^93,94^ CD244,^95^ and TNF superfamily members OPG and TWEAK.^96^ Atomoxetine treatment was associated with reduced levels of the pro-inflammatory cytokines CDCP1, CD244, and TWEAK. Interestingly, reduced expression of CDCP1 is protective in experimental autoimmune encephalomyelitis,^97^ reduced expression of CD244 improves survival in sepsis,^98^ and reduced TWEAK activation is protective in a variety of models of inflammation, fibrosis, and angiogenesis.^99,100^ Atomoxetine increased levels of osteoprotegerin (OPG), a decoy receptor for TNF receptor, with multiple functions and effects including promotion of vascular health and cell survival.^101^ Hence, although roles of these proteins in brain is limited and effects of atomoxetine need further study, these results are generally consistent with a potential therapeutic benefit. Another LC function is regulation of neurotrophin signaling and neurogenesis,^8,41,44^ and in our study atomoxetine increased plasma BDNF levels. Thus, our proteomics approaches provide independent evidence that atomoxetine modulated several pathophysiological processes important in Alzheimer’s disease, including synaptic function, metabolism, neuroinflammation, and neurotrophin signaling.

There are several limitations of our study. Although the cross-over study design allows each participant to serve as their own control, thus increasing power, a weakness in this approach is the carry-over effects that we observed. Because atomoxetine has a short drug half-life (∼5 h)^56^, the carry-over effects observed in the FDG-PET analyses suggests longer term effects of treatment. Hence, future trials with atomoxetine would benefit from a parallel arm design to better address the potentially disease-modifying effects. Our study was also limited in that it involved only a single site, enrolled a modest number of participants, and was relatively short (6 months of treatment) for evaluation of disease-modifying effects. Although the study successfully achieved the primary outcomes of safety and being well tolerated in individuals with MCI, the other primary outcome assessing CSF levels of two preselected inflammatory analytes, IL1- and TECK, suffered from methodological limitations and were undetectable in most samples. We circumvented this limitation with broad exploration of other analytes using novel approaches such as mass spectrometry and PEA assays that were not available during the planning of the study. Although promising, these results should be considered as exploratory given the large number of analytes measured.

Given that increased CSF levels of NE and its metabolites are found in Alzheimer’s disease and correlate with pathology and cognitive decline,^102,103^ it might seem counterintuitive to treat the disease with a NET inhibitor that further increases NE transmission. There is evidence for LC hyperactivity in surviving neurons during the degenerative process, resulting in a complex dysregulation of NE transmission.^7,104,105^ Importantly, atomoxetine does not just indiscriminately increase synaptic NE levels; it also improves the phasic-to-tonic ratio of LC firing,^52^ which is associated with focused attention important for some aspects of learning and memory. Thus, atomoxetine may ameliorate neuroinflammation by increasing NE, while simultaneously improving cognition and behavior by “normalizing” LC activity in the face of pathology and degeneration. More preclinical and clinical research will be required to identify the mechanistic relationships between LC degeneration, NE and metabolite levels, and treatment efficacy.

In sum, this phase II study of atomoxetine demonstrated excellent safety, tolerability, and target engagement in individuals with MCI, all advantages of repurposing a well-studied FDA-approved medication. The study also provided evidence supporting potential disease-modifying effects of atomoxetine on a variety of CSF markers (Tau, pTau181, brain-linked mass spectrometry proteomic panels, inflammatory analytes measured by PEA assay) and imaging markers (FDG-PET and rsfMRI). Given the failure of disease-modifying therapies to date, the results warrant consideration of future clinical trials of atomoxetine to enhance LC function for disease-modification.

## Abbreviations

Aβ: amyloid-beta
ADAS-Cog: Alzheimer’s Disease Assessment Scale-Cognitive Subscale
ADDF: Alzheimer Drug Discovery Foundation
ADNI: Alzheimer’s disease neuroimaging initiative
BDNF: brain-derived neurotrophic factor
CSF: cerebrospinal fluid
DHPG: 3,4-dihydroxyphenylglycol
DREADD: designer receptors exclusively activated by designer drugs
DSMB: Data and Safety Monitoring Board
FDG: fluorodeoxyglucose
GDS: Geriatric Depression Scale
LC: locus coeruleus
L-DOPS: L-3,4-dihydroxyphenylserine
LM: Logical Memory
MCI: mild cognitive impairment
MMSE: Mini Mental Status Exam
MS: mass spectrometry
NE: norepinephrine
NET: norepinephrine transporter
TECK: Thymus Expressed Chemokine
TMT: tandem mass tagging

## Acknowledgements

The authors thank the staff of the Emory Goizueta Alzheimer’s Disease Research Center, with special gratitude to CeeCee Manzanares, Margaret Walker, and Janet Cellar for their operational support, and to the participants and their families. We also thank Patti Sullivan for conducting the catechol assays.

## Funding

Funding for the study was provided by the Cox and Kenan Family foundations, and the Alzheimer’s Drug Discovery Foundation. The research reported here was also supported (in part) by the Division of Intramural Research, NIH, NINDS.

## Competing interests

The authors report no competing interests

## Supplementary material

Supplementary material is available at *Brain* online

## Notes

### Competing Interest Statement

The authors have declared no competing interest.

### Clinical Trial

NCT01522404

### Funding Statement

Funding for the study was provided by the Cox and Kenan Family foundations, and the Alzheimers Drug Discovery Foundation. The research reported here was also supported (in part) by the Division of Intramural Research, NIH, NINDS.

### Author Declarations

The trial was registered with ClinicalTrial.gov (NCT01522404) after the protocol and informed consent form were reviewed and approved by the Emory Institutional Review Board (IRB) committee (IRB00054397).

