## Supplementary Material for "A Phase II Study Repurposing Atomoxetine for Neuroprotection in Mild Cognitive Impairment"

**Supplementary Table 1. Comparison of Adverse Events Between Atomoxetine and Placebo Treatments**

| **Symptom** | **Atomoxetine (n=39)** | **Placebo (n=37)** |
| --- | --- | --- |
| Serious Adverse Event, n (%) |  |  |
| Fractured right pelvis | 1 (2.6) | 0 |
| Acute appendicitis | 0 | 1 (2.7) |
| Hypothermia/dizziness | 1 (2.6) | 0 |
| Severe Elevated Blood Pressure | 0 | 1 (2.7) |
| Dysautonomia | 1 (2.6) | 0 |
| Headache Pain | 0 | 1 (2.7) |
| Non-Serious Adverse Event, n (%) |  |  |
| Gastrointestinal symptoms | 12 (30.8) | 4 (10.8) |
| Dry mouth | 10 (25.6) | 2 (5.4) |
| Dizziness | 10 (25.6) | 8 (21.6) |
| Falls | 6 (15.4) | 2 (5.4) |
| Upper respiratory infection | 6 (15.4) | 9 (24.3) |
| Headache | 5 (12.8) | 2 (5.4) |
| Constipation | 4 (10.3) | 1 (2.7) |
| Suicide | 3 (7.7) | 0 |
| Tremor | 3 (7.7) | 0 |
| Agitation | 3 (7.7) | 1 (2.7) |
| Loss of appetite | 3 (7.7) | 1 (2.7) |
| Fatigue | 3 (7.7) | 3 (8.1) |
| Fracture | 2 (5.1) | 0 |
| Loss of Weigh | 2 (5.1) | 0 |
| Anxiety | 2 (5.1) | 1 (2.7) |
| Injury | 2 (5.1) | 1 (2.7) |
| Rash | 2 (5.1) | 2 (5.4) |
| Hypokalemia | 1 (2.6) | 2 (5.4) |
| Pre-Cancerous | 1 (2.6) | 2 (5.4) |
| Back pain | 1 (2.6) | 3 (8.1) |
| Cancer | 1 (2.6) | 3 (8.1) |
| Cramp | 0 | 2 (5.4) |

*Only the Adverse Events above 5% threshold are listed under the Non-Serious Adverse Event.

**Supplementary Table 2. Comparison of Change in Blood Pressure and Heart Rate between two Treatment Arms**

| **Variable** | **Visit** | **Active/Placebo**  **Median (P25 - P75) (n)** | **Placebo/Active**  **Median (P25 - P75) (n)** | **p value** |
| --- | --- | --- | --- | --- |
| Change in systolic BP from baseline | 6 month | 5.5 (-9.0, 12.0) (18) | 4.0 (-6.0, 15.0) (19) | 0.86 |
|  | 12 month | -4.0 (-10, 10.0) (17) | 3.0 (-7.0, 19.0) (19) | 0.25 |
| % Change in systolic BP from baseline | 6 month | 4.3 (-6.5, 7.6) (18) | 2.6 (-4.6, 12.8) (19) | 0.94 |
|  | 12 month | -3.2 (-7.6, 6.8) (17) | 3.0 (-5.3, 14.6) (19) | 0.21 |
| Change in diastolic BP from baseline | 6 month | 3.5 (-5.0, 6.0) (18) | 1.0 (-5.0, 11.0) (19) | 0.81 |
|  | 12 month | -4.0 (-9.0, 2.0) (17) | -3.0 (-8.0, 7.0) (19) | 0.22 |
| % Change in diastolic BP from baseline | 6 month | 4.4 (-6.2, 8.3) (18) | 1.4 (-6.2, 16.4) (19) | 0.83 |
|  | 12 month | -4.9 (-11, 3.1) (17) | -4.1 (-12, 9.2) (19) | 0.27 |
| Change in heart rate from baseline | 6 month | 9.0 (6.0, 12.0) (17) | 1.0 (-4.0, 5.0) (19) | 0.0006 |
|  | 12 month | 4.0 (-2.0, 8.0) (17) | 8.0 (5.0, 12.0) (19) | 0.012 |
| % Change in heart rate from baseline | 6 month | 14.8 (10.3, 19.4) (17) | 2.2 (-6.5, 8.6) (19) | 0.0006 |
|  | 12 month | 6.9 (-3.4, 10.5) (17) | 13.8 (8.7, 24.5) (19) | 0.0107 |
| Change in weight from baseline | 6 month | -4.0 (-15, 0.0) (18) | -1.0 (-3.0, 1.0) (19) | 0.07 |
|  | 12 month | -1.0 (-6.0, 3.0) (17) | -4.0 (-7.0, -1.0) (19) | 0.25 |
| % Change in weight from baseline | 6 month | -3.0 (-7.0, 0.0) (18) | -0.8 (-2.0, 0.6) (19) | 0.07 |
|  | 12 month | -0.9 (-3.3, 2.3) (17) | -2.3 (-4.8, -0.8) (19) | 0.24 |

**Supplementary Figure 1. Adjusted Effect of Treatment of Atomoxetine vs Placebo on Neuropsychological Measures.** Values are estimated differences and corresponding 95% confidence interval. The asterisked results for ADAS13 and Trails B were raw p values of *p<0.05, but were not significant after adjusting for multiple comparisons.

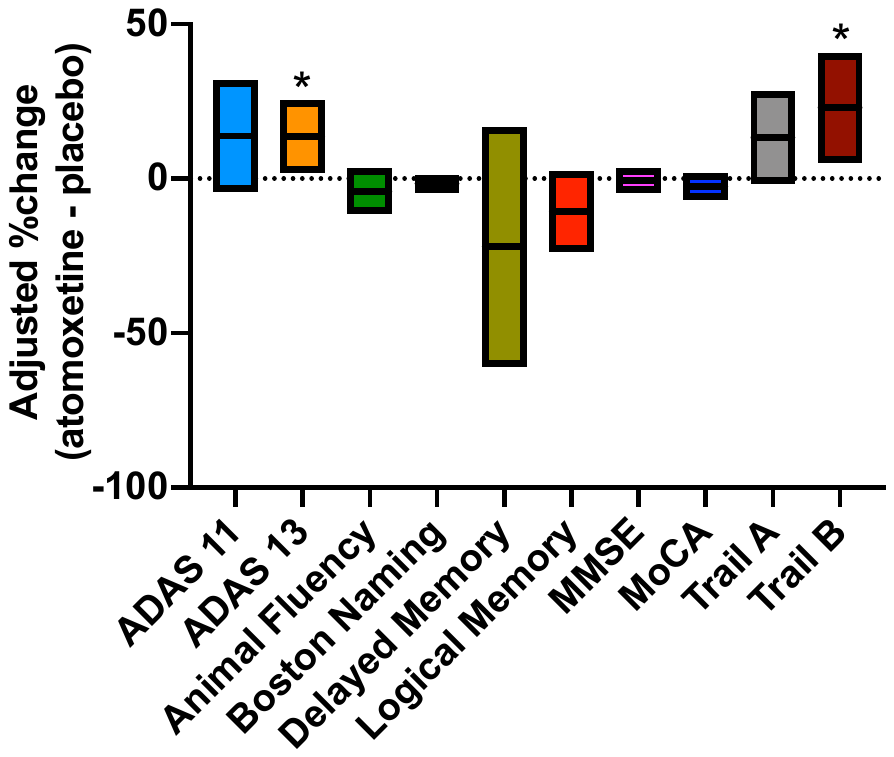

**Supplementary Figure 2. Adjusted Effect of Treatment of Atomoxetine vs Placebo on Absolute Change in CSF Biomarkers of Inflammation**. A) Immunoassays for targeted biomarkers shown. B) Proximity Extension Assays using Olink Inflammation panel. Only the significantly changed biomarkers are shown, from a panel of 92 analytes. Values are estimated differences and corresponding 95% confidence interval. * p<0.05; **p<0.01

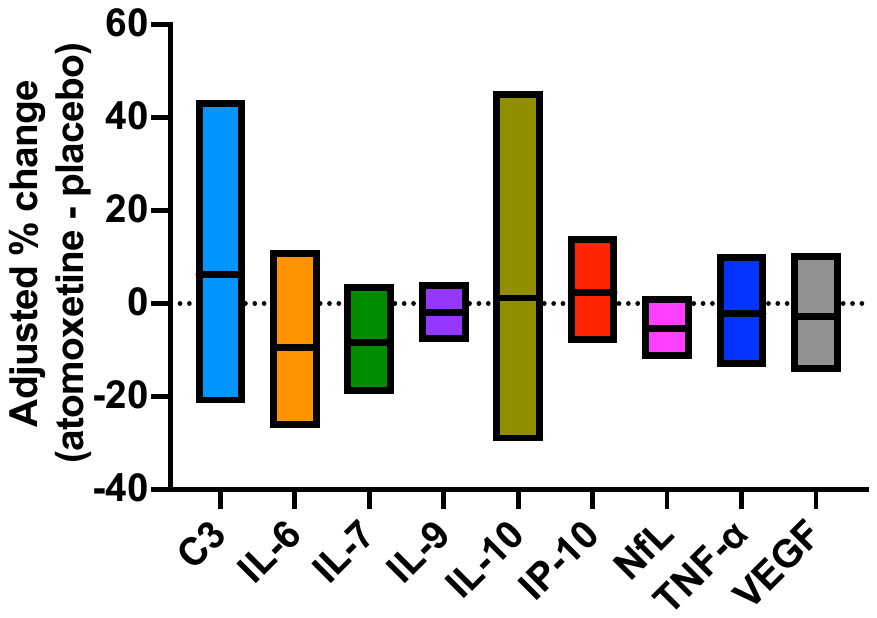

A

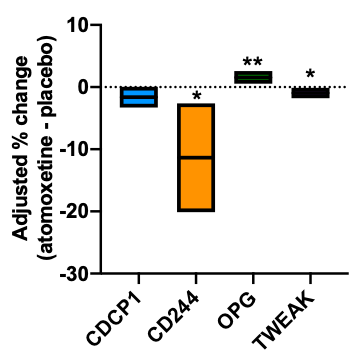

B

**Supplementary Figure 3. Adjusted Effect of Treatment of Atomoxetine vs Placebo on Absolute Change in Plasma Biomarkers.** A) Luminex assay results for targeted CSF biomarkers shown (performed by Dr. William Hu). B) Mesoscale multi-plexed immune-assay results for plasma analytes (performed by Dr. Malú G. Tansey). Values are estimated differences and corresponding 95% confidence interval. * p<0.05

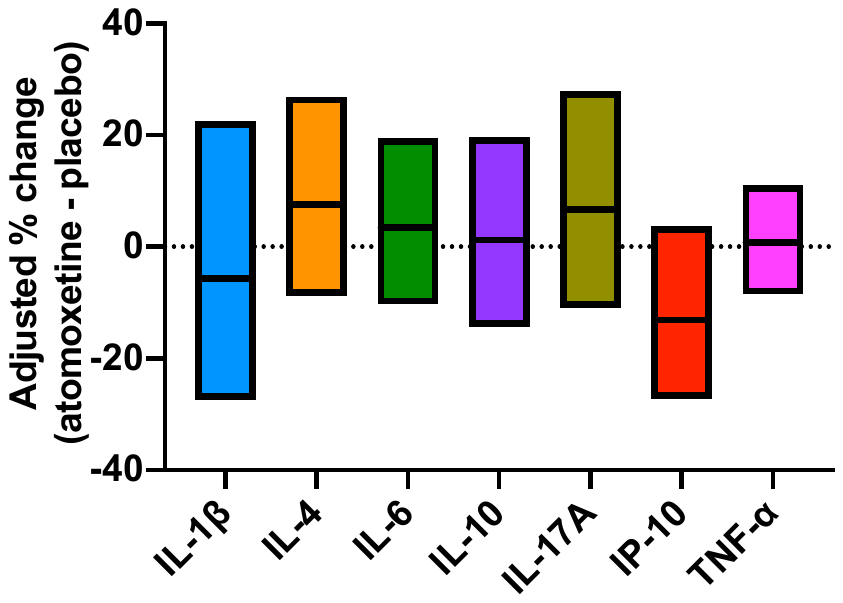

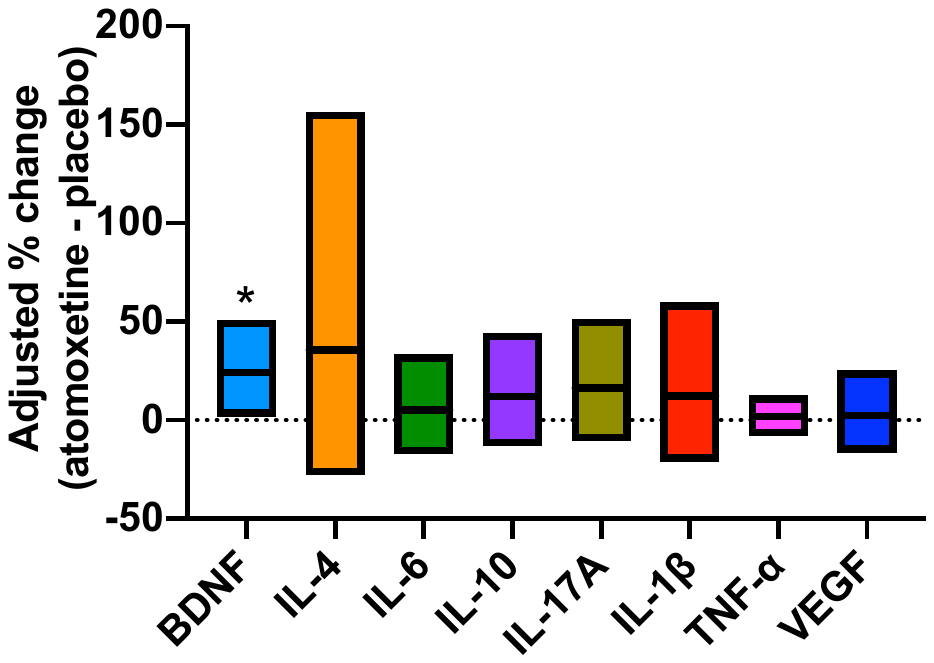
